# Contributions to palliative and end-of-life care by community health nursing services: improving care through national and regional service evaluations

**DOI:** 10.64898/2026.04.21.26351027

**Authors:** Sophie Pask, Assem Khamis, Tina Jarrett, Joanna M. Davies, Catherine J. Evans, Fliss E. M. Murtagh

## Abstract

**Aim(s):** To describe adult palliative and end-of-life care provision by community health nursing services using a:

- National dataset (2013-2024) to report patterns in service provision over time.
- Regional dataset (2022/23, 2023/24 and 2024/25) to describe palliative and end-of-life care activities.

**Design:** Secondary analyses of existing national and regional datasets.

**Methods:** We used national data to describe the populations served; workforce; referrals; unique service users seen annually; contacts; time on caseload; care delivered/care locations; support to other teams/processes; and deferred care. Regional data was used to examine palliative and end-of-life care activities in the context of all nursing care delivered.

**Results:** Nationally, referrals to community health nursing services increased steadily from 4,000 to 6,000 per 100,000 weighted population between 2013 and 2024, while unique service users remained stable (around 2,600–2,800). Median average time on caseload reduced markedly from over 150 days to around 50 days, despite stable contact frequency (median 23 total contacts per service user) and duration (median 26 minutes for face-to-face contacts).

Regional data showed that palliative and end-of-life care consistently accounted for 9.6% of all community nursing clinical time (30–32 hours per 1,000 population annually) across three years, even as total care hours declined. A disproportionate amount of palliative and end-of-life care occurred out-of-hours.

**Conclusion:** Increasing referrals and shorter time on caseloads indicate a system under pressure. Time spent on palliative and end-of-life care by community health nursing teams has remained stable over time, despite growing population need. Workforce capacity, skill mix, and out-of-hours provision need to align to support high-quality, person-centred care in the community.

**Implications for the profession and/or patient care:** This evidence informs better planning to ensure sufficient provision and workforce in community health nursing.

**Patient and public contribution:** Patients, family carers and public members contributed to interpreting findings and implications for practice.

**What does this paper contribute to the wider global clinical community?:**

- Provides combined national and regional data to describe the scale and nature of palliative and end-of-life care delivered by community health nursing services over time.
- Findings give a detailed picture of how community health nursing services are under pressure because of increasing referrals and being required to deliver a greater breadth of tasks.
- Models of community health nursing are changing with shorter care episodes and significant palliative and end-of-life care workload (with distinctive challenges out-of-hours).

## 1. Introduction

Community health nurses and their wider team (e.g. healthcare assistants and nursing associates) are pivotal to the delivery of holistic person-centred care in the community, including supporting people with palliative and end-of-life care needs.^1–3^ In England, ‘community health nursing’ includes district and community nurses, and their wider team, for whom palliative and end-of-life care provision is one part of the nursing care they provide across the lifespan.^4^ Palliative care aims to improve the quality of life of people and their families facing advanced illness through a holistic approach (including as early identification, comprehensive assessment, and symptom management).^5^ End-of-life care is a key part of this, focusing on helping people to live as well as possible in the last phase of life.^5^

Population ageing and the growing prevalence of long-term health conditions mean that more people will need palliative and end-of-life care over the next 15 years, especially in the community setting.^6,7^ At the same time, changes in multidisciplinary team working (e.g. more remote working by general practitioners), workforce challenges, and resource constraints have reshaped the landscape of community health nursing in the UK.^4,8–11^ All of which has implications for team structure, skill mix, workload and caseload complexity and consequently the delivery of palliative and end-of-life care.^4,8,9,11^

## 2. Background

Community health nursing teams often take the lead in palliative and end-of-life care within the wider multi-professional team, and their roles have extended in recent years.^8^ Despite the centrality of palliative and end-of-life care in community health nursing, data on its scale and nature remain limited. Estimates (both historical and contemporary) of the number of palliative and end-of-life care patients on caseloads and the clinical time spent with these patients in the UK vary widely.^12–14^ More recently, an NHS Benchmarking Network (2020) report indicated that 6% of community health nursing clinical time is spent on palliative care.^13^ A nationally representative survey of England and Wales showed that 53.4% of people dying from expected causes of death had contact with a community health nurse in the last 3 months of life (with a median of 5 contacts, interquartile range: 2-10).^14^ UK survey data showed that 1,156 community health nurses self-reported spending 23.5% of their most recent shift providing end-of-life care.^15^ Of all 1,471 nurses surveyed (including 174 from specialist palliative care services), 11.6% indicated that they had to defer end-of-life care visits during their most recent shift due to staff shortages, demand exceeding capacity and other systemic barriers.^15^ In addition, 52% of respondents who provided end-of-life care during their last shift reported being unable to deliver certain aspects of care to professionally acceptable standards.^15^

These changes in patterns of nursing care are occurring alongside policy ambitions to shift care from hospital to the community,^16,17^ and the development of a Modern Service Framework for palliative and end-of-life care to support long-term planning, sustainable investment and consistent delivery of high-quality evidence-based care.^18^ Understanding and quantifying service demand and how community health nursing teams spend their time in relation to palliative and end-of-life care is essential for guiding practice, capacity modelling, workforce planning (including team composition), and quality improvement.

## 3. The Study

The aim of this work is to describe provision of adult palliative and end-of-life care by community health nursing services using:

- The NHS Benchmarking Network national dataset (2013-2024) from England to report patterns in service provision over time.
- The regional ‘Care in Focus’ dataset (2022/23, 2023/24 and 2024/25) from Birmingham to describe the nature, and time spent on, palliative and end-of-life care activities within the context of all general nursing care provided.

## 4. Methods

### 4.1 Design

Secondary analyses of existing anonymised clinical data conducted as national and regional service evaluations. Service evaluations are conducted to define the care provided within an existing service.^19^ The reporting of this work was guided by the Revised Standards for Quality Improvement Reporting Excellence (SQUIRE 2.0).^20^

### 4.2 Description of data sources (population, setting and data variables)

#### 4.2.1 The NHS Benchmarking Network national dataset

The NHS Benchmarking Network is a UK-wide, impartial, member-led, community of health and care organisations. Annual surveys (submissions) are received from participating provider organisations at organisational and site/service level across the UK. Community health nursing data has been benchmarked annually for over 10 years in the Community Services Project, and more recently, as a standalone project.^21,22^ For this paper, we used the annual submissions from participating organisations in England between 2013 and 2024.^23,24^ The services that responded each year varied (see Table 1 in Supplementary File 1).

**Table 1.** Locations in which their service is provided (n= 65)

| Locations in which service is provided in... | Yes n (%) | No n (%) | Missing n (%) |
| --- | --- | --- | --- |
| Own home<sup>a</sup> | 64 (98%) | 0 (0%) | 1 (2%) |
| Care home<sup>b</sup> | 64 (98%) | 0 (0%) | 1 (2%) |
| Outpatient<sup>c</sup> | 54 (83%) | 9 (14%) | 2 (3%) |
| Acute hospital<sup>d</sup> | 22 (34%) | 41 (63%) | 2 (3%) |
| Community and day hospital<sup>e</sup> | 15 (23%) | 48 (74%) | 2 (3%) |
| Hospices | 8 (12%) | 55 (85%) | 2 (3%) |
| Community facilities and other<sup>f</sup> | 24 (37%) | 39 (60%) | 2 (3%) |

a Refers to service user's own home (including hostels and supported living facilities).
b Refers to nursing and residential homes.
c Refers to clinics and health centres, day centres and GP surgeries.
d Refers to acute hospitals and mental health inpatient facilities.
e Refers to community hospital and day hospital.
f Refers to community facilities (e.g. community halls and centres), education facilities, leisure facilities, specialist schools, the justice system (including prisons) and 'other'.

In this dataset, community health nursing services were defined as providing nursing care in a particular geographical area (frequently in the patients’ own home or in residential care homes). Core functions include a holistic assessment of patients’ needs, referrals to social care and for equipment, prescription of medication or referrals for medication, continence care, follow-up care for patients discharged from hospital, long-term care for chronically ill patients, end-of-life care, wound care and support for families and carers. Out-of-hours and rapid response community health nursing is usually provided, as well as community matrons, healthcare assistants and nursing associates that operate as part of the community health nursing team. Dedicated rapid response teams and specialist nursing services (e.g. condition specific services) are not included.

Available data included population served (including weighted population where base populations are weighted by age, sex and GP practices needs index (including deprivation)), workforce (in establishment – budgeted positions (including occupied and vacant roles) and in post – actual number of employees occupying roles), referrals (received, accepted and source), unique service users seen in the year, contacts (face-to-face, non-face-to-face and total), average time on caseload in days, types of care provided, locations care is provided in, support to other teams/processes, location of patients when discharged from their service, and deferred care. Further detail on the metrics used and how we have defined or re-grouped them are available in Supplementary File 1.

#### 4.2.2 Care in Focus regional dataset

‘Care in Focus’ is a clinically led initiative by Birmingham Community Healthcare NHS Foundation Trust. Data is collated annually from 27 community health nursing teams (two of which cover the out-of-hours period) in Birmingham, which sits within the wider Birmingham and Solihull Integrated Care System. During normal working hours (8am-6pm), teams work in five clusters comprising North, East, West, South and Central localities. Each locality has four planned care teams and one unplanned care team. All localities are then covered by an evening service team (6pm-10pm) and a night service team (9:30pm-8am) in the out-of-hours period. The evening service is a ‘bridging’ service and provides about 40% of planned evening visits and about 60% of on call (unplanned) visits, whilst the night service is 100% on call (unplanned visits only). Detail on team structure can be found in Figure 1 in Supplementary File 1.

**Figure 1.**
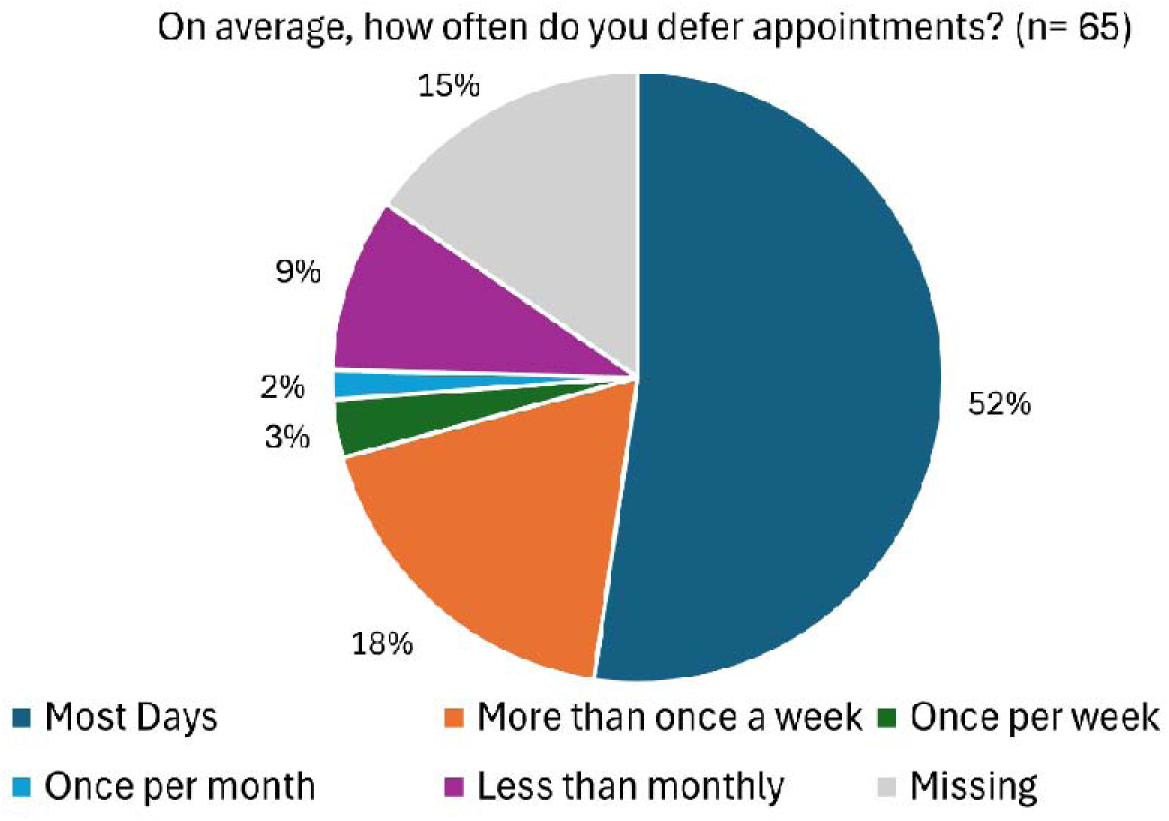
How often services defer appointments on average

The ‘Care in Focus’ tool intends to capture all activity (both planned and urgent/unplanned) but is largely reflective of planned care. Planned data is collected from the electronic patient record (Rio) and emergent visits are directly recorded in the tool. It captures the time spent on nursing activities, including detailed categorisation of these activities into type and duration. The tool is used to provide a framework the allocation of workload and inform team structure. In its development, all clinical interventions were considered and included in terms of average length of time, skill, and banding required and compared to historic electronic data. We sought recent data for a period of three years on all palliative and end-of-life care activities, in the context of all care provided, by selecting all activities that related to palliative and end-of-life care from a complete list of all community health nursing activities (see Table 2 in Supplementary File 1 for selected activities and how these were grouped).^25^ Generic activities which might have been delivered towards the end of life are not included; therefore, our estimates will be conservative.

### 4.3 Data management and analysis

Data were managed and analysed using Microsoft Excel. Descriptive analyses (such as frequencies, median and interquartile range) were used to describe variables.

For national data, submissions are organisation-specific and year-specific, and it is not possible to track the same organisations across different years. Any changes between years seen in the data may be a result of this sample variation, although the high proportion of services included each year among all England services mitigates against this. Missing data were present across years and within variables, leading to variation in the sample size ‘N’. We have therefore transparently reported the relevant ‘N’ for each analysis.

### 4.4 Patient and Public Involvement

We held an online session with our established Patient and Public Involvement Group (including patients, family carers and members of the public) on 7^th^ January 2026. The purpose of this meeting was to share the analysis and seek their views on and interpretations of the data. The group discussion helped us to shape how we presented and prioritised the data, and to consider how key findings from this service-level data relate to issues and concerns that matter to people affected.

### 4.5 Ethical Considerations

As these service evaluations used routinely collected data and there was no change in patient care, formal ethical approval was not required as per local governance policies. Data represent services and were anonymised prior to receipt of the data. The NHS Benchmarking Network Delivery Team (hosted by East of England Community Health and Care NHS Trust, United Kingdom) confirmed that the data must be referenced but formal ethical approval was not required. The Birmingham Community Healthcare NHS Foundation Trust Research and Innovation Team (Birmingham, United Kingdom) reviewed the project and confirmed that a formal review or issuing approvals were not required.

## 5. Findings

The results are presented in three parts:

5.1 National cross-sectional analysis for 2024

5.2 National repeated cross-sectional analyses (for 2013 to 2024)

5.3 Regional data, with repeated cross-sectional analyses (for 2022/23, 2023/24 and 2024/25)

### 5.1 National cross-sectional analysis for 2024

In 2024, responses to the annual NHS Benchmarking Network survey were received from 65 services (from to 48 Trusts) who participated across England.

#### 5.1.1 Population served by community health nursing services

Of the 65 services, 64 provided data on the population served. These 64 services reported serving a total population of 30,641,362. Therefore, based on mid-year 2024 population estimates for England by the Office for National Statistics,^26^ the population served in this data accounts for 52% (30,641,362/58,620,101) of the total population in England. The median population served by each service is 376,629 (IQR 234,039 – 651,766), with a minimum of 140,430 and a maximum of 1,182,804. 63 services provided data on the proportion of people aged ≥65, with data missing from two services. A total of 5,674,595 people were aged ≥65, which is 18.7% of the total population served (n= 30,378,523 for the 63 services). This mirrors the proportion of people aged ≥65 in the total population of England in ONS data.^26^ 62 of the 65 services provided weighted population data, serving a total weighted population of 29,570,331. The median weighted population served is 382,301 people (IQR 208,581 – 681,657).

#### 5.1.2 Workforce (in establishment and in post) Registered nurses

For 59 services with available data, the median whole time equivalent in establishment per 100,000 weighted population was 32 (IQR 26 – 42). The median whole time equivalent in post per 100,000 weighted population was 28 (IQR 21 – 40).

##### Nursing support staff

For 59 services with available data, the median whole time equivalent in establishment per 100,000 weighted population was 11 (IQR 9 – 14). The median whole time equivalent in post per 100,000 weighted population was 10 (IQR 8 – 13).

#### 5.1.3 Referrals Referrals received

63 services provided data on the number of referrals received. The median number of referrals received by each service in 2024 was 22,090 (IQR 11,903 – 38,699). For the 60 services where weighted population data was available, the median number of referrals received by each service per 100,000 weighted population was 6,157 (IQR 4,474 – 8,757), with a minimum of 1,131 and a maximum of 23,073.

##### Referrals accepted

61 services provided data on the number of referrals accepted in 2024. The median number of referrals accepted was 20,885 (IQR 11,693 – 34,766). For the 59 services where weighted population data was available, the median number of referrals accepted by each service per 100,000 weighted population was 5,630 (IQR 4,068 – 7,844), with a minimum of 1,103 and a maximum of 23,030.

##### Referrals not accepted

For the 61 services that reported both referrals received and referrals accepted, the median percentage of referrals not accepted was 3% (IQR 0% – 7%), with the highest percentage of referrals not accepted from those received being 29% in one service.

##### Sources of referrals

Community nursing services were asked to provide a breakdown of 17 possible sources of referrals, although the data provided varied by service (see Figure 2 in Supplementary File 1). The main source of referrals to the community health nursing services were from general practice at 37% (with 60 services providing data).

**Figure 2.**
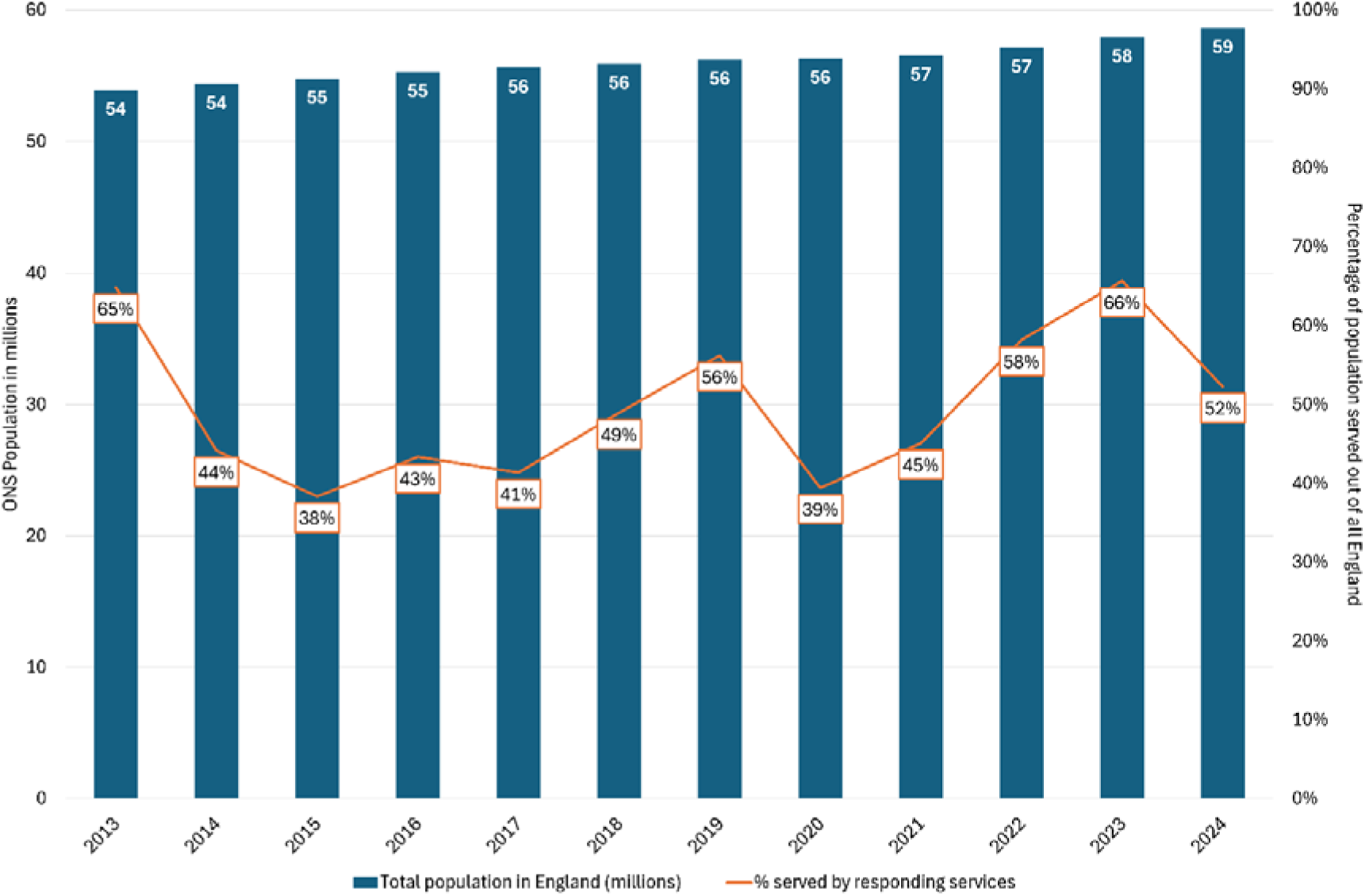
Percentage of the population served by community health nursing teams in England between 2013 and 2024

#### 5.1.4 Number of unique services users seen in the year

59 of 65 services reported on the number of unique service users. The median number of unique service users seen in the year was 10,618 (IQR 7,730 – 15,903). For the 57 services where data was available, the median number of unique service users seen in the year by each service per 100,000 weighted population was 2,716 (IQR 2,292 – 3,926).

#### 5.1.5 Contacts Face-to-face

61 services provided data on both face-to-face contacts and referrals accepted. The median face-to-face contacts per accepted referral by service was 11 (IQR 8 – 14). In considering face-to-face contacts by unique service users (n= 59), the median face-to-face contacts per unique service user by service was 20 (IQR 16 – 28).

##### Non-face-to-face

60 services provided data on both non-face-to-face contacts and referrals accepted. The median non-face-to-face contacts per accepted referral by service was 0.8 (IQR 0.3 – 2). In considering non-face-to-face contacts by unique service users (n= 59), the median non-face-to-face contacts per unique service user by service was 1.6 (IQR 0.6 – 3.7).

##### Total contacts

Total contacts were also considered by accepted referral (n= 61) and unique service users (n= 59), respectively. The median number of total contacts per accepted referral by service was 12 (IQR 9 – 16). The median number of total contacts per unique service user by service was 23 (IQR 18 – 29).

##### Average length of contacts

52 services provided data on the average length of contacts. The median length of face-to-face contacts was 26 minutes (IQR 20 – 34), with a minimum of 13 and a maximum of 57 minutes. The median length of non-face-to-face contacts was 12 minutes (IQR 9 – 18), with a minimum of 4 and a maximum of 39 minutes.

#### 5.1.6 Average time on caseload

50 services provided data regarding the average time on caseload. The median average time on caseload for service users discharged was 51 (IQR 35 – 79) calendar days, with a minimum of 16 and a maximum of 312 calendar days.

#### 5.1.7 Locations care is provided in

Services reported the various locations in which they provided their service, see Table 1 for the breakdown. Some community health nursing services provided ‘in-reach’ into hospitals and other health facilities, according to their localities. Some services also provided support to other teams and processes, which is presented in Figure 3 in Supplementary File 1.

**Figure 3.**
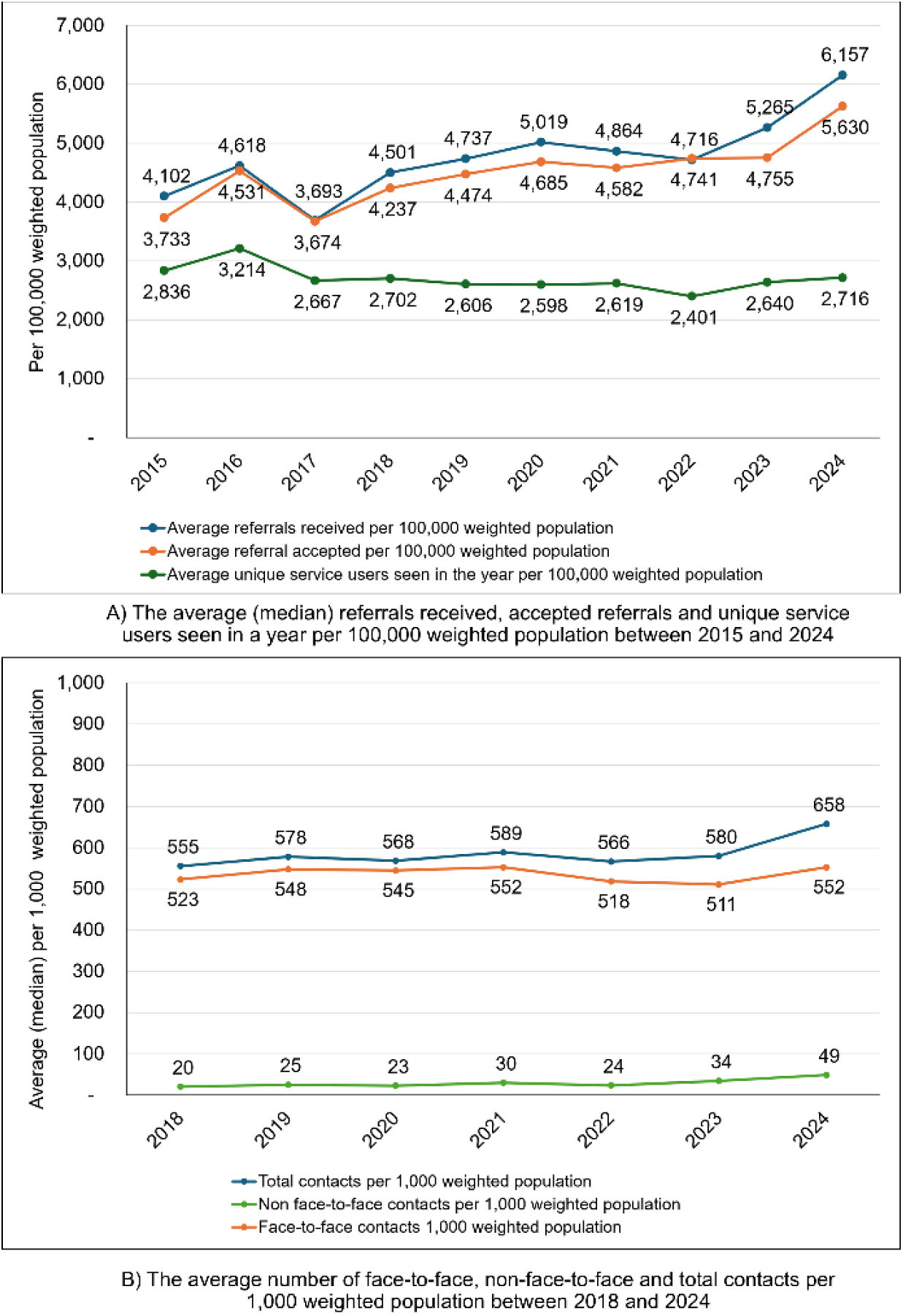
Referrals (received and accepted) and unique service users seen in the year, and contacts by weighted population Note: the ‘average’ in the repeated cross-sectional analyses refers to the median statistics across all submissions received in each year/in that specific year.

#### 5.1.8 Deferred care

Figure 1 below shows how often services deferred appointments for all care delivered (i.e. a planned visit postponed due to insufficient staff capacity to meet demand and the need to prioritise patients), with 52% of services deferring appointments most days.

### 5.2 National repeated cross-sectional analyses (for 2013 to 2024)

#### 5.2.1 Population served

Figure 2 presents the total population of England (using Office for National Statistics data)^26^ between 2013 and 2024 and the percentage of the unweighted population served by services responding to the annual benchmarking survey.

The percentage of people aged ≥65 in the population served by community health nursing services increased from 13% in 2013 to 19% in 2024.

#### 5.2.2 Referrals, unique service users seen in the year and contacts by weighted population

Weighted data was only available from 2015 for referrals (received and accepted) and unique service users seen in the year and contact data was only available from 2018. The median number of referrals received, accepted referrals and unique services users seen in a year per 100,000 weighted population between 2015 and 2024 are presented in the figure below (A). Referrals received and accepted have increased over time, whilst the number of unique service users seen in the year has remained stable over the years. The figure also shows the average number of total contacts per 1,000 weighted population has largely remained stable (B), but with a slight increase in recent years.

#### 5.2.3 In-depth analysis of contacts (by accepted referrals and unique service users seen in the year), length of contacts and time on caseload

Note: the ‘average’ in the repeated cross-sectional analyses refers to the median statistics across all submissions received in each year/in that specific year.

Figure 4 presents contacts (face-to-face, non-face-to-face and total) by accepted referrals (A) and unique service users seen the year (B), as well as the average length of contacts in minutes (C) and average time on caseload (D).

**Figure 4.**
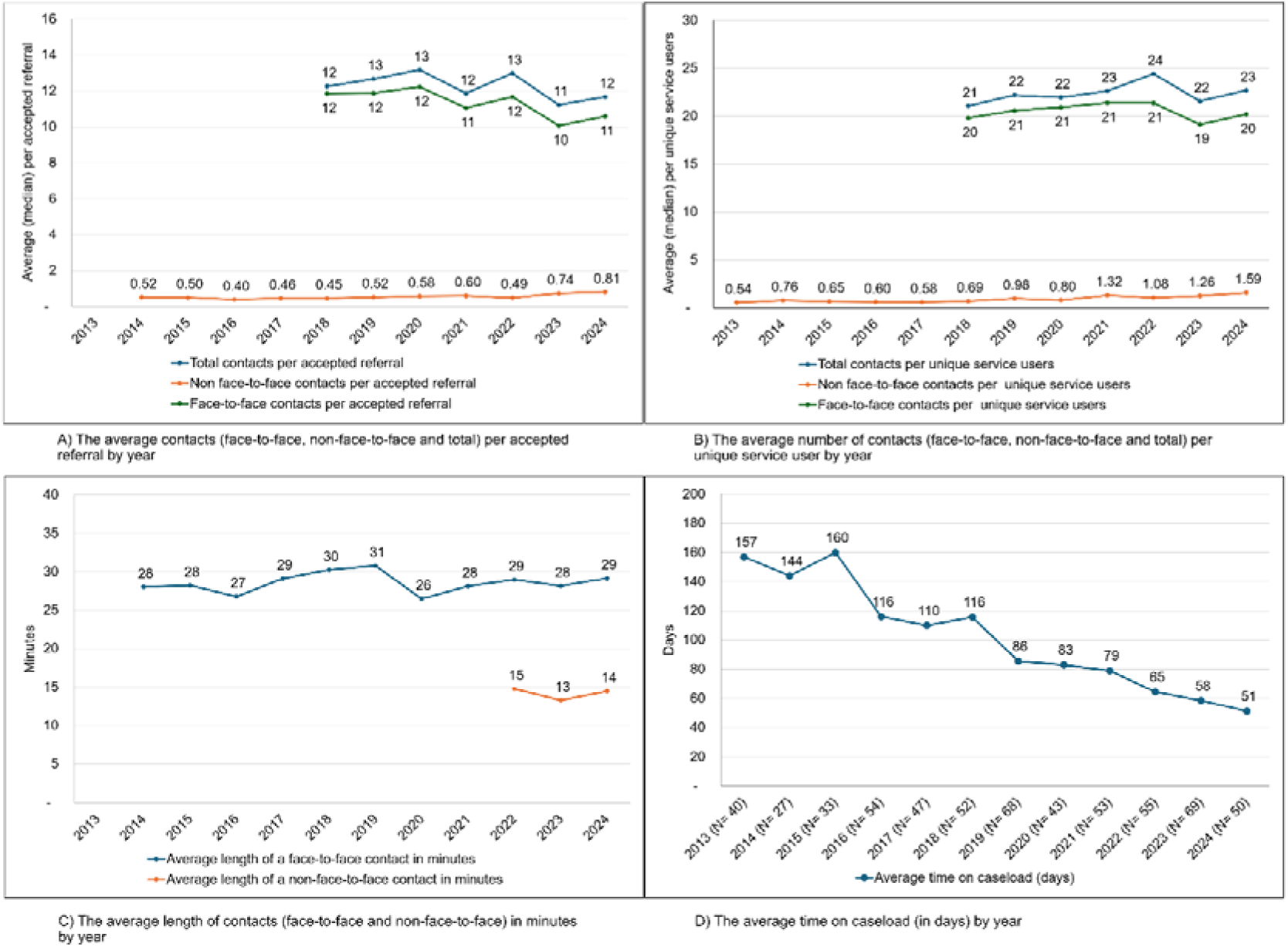
Contacts (by accepted referrals and unique service users seen in the year), average length of contact and time on caseload over time between 2013 and 2024 Note: the ‘average’ in the repeated cross-sectional analyses refers to the median statistics across all submissions received in each year/in that specific year.

The average number of contacts (face-to-face, non-face-to-face and total) per accepted referral and per unique service user have remained stable, although full data are only available from 2018 onwards for face-to-face and total contacts.

The average length of face-to-face contacts in minutes has remained steady. Whilst the average time on caseload (in days) has decreased substantially between 2013 and 2024.

#### 5.2.4 Types of care provided

Figure 5 summarises the types of care provided by community health nursing services (from a predefined list in the annual survey) between 2017 and 2024, some of which are not exclusive to but were considered to relate to palliative and end-of-life care. Generally, these types of care appear to be increasingly offered by more community health nursing services.

**Figure 5.**
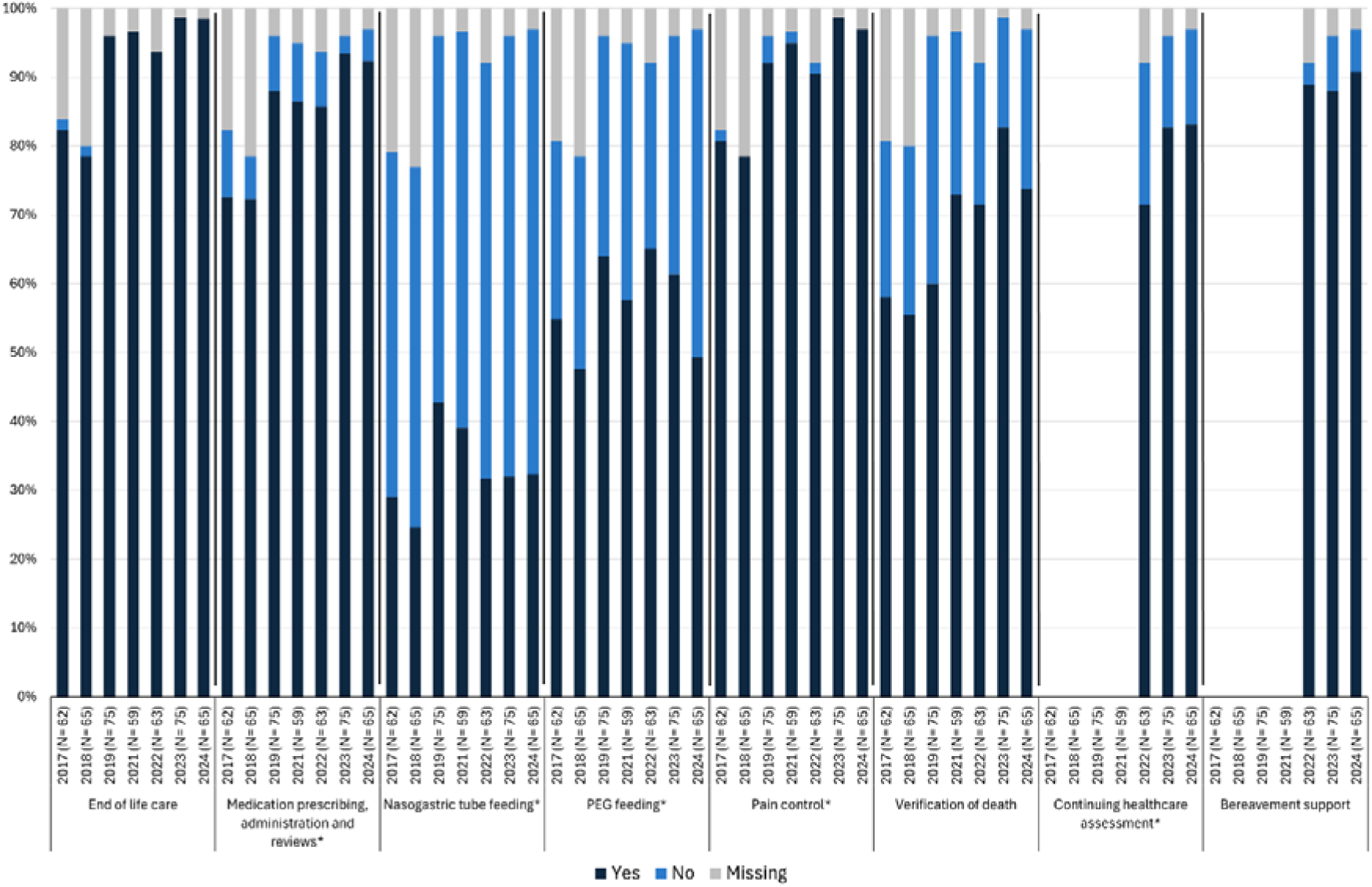
Types of care provided by services by year Note: PEG Percutaneous endoscopic gastrostomy. * These types of care are not all exclusive to palliative and end-of-life care and may be provided as part of general nursing care. Continuing healthcare assessment and bereavement support only reported from 2022.

### 5.3 Regional data, with repeated cross-sectional analyses (for 2022/23, 2023/24 and 2024/25)

#### 5.3.1 Population served

Using 2021 Census data,^27^ the population for the Birmingham area is 1,144,923, with 51.1% female; 13.1% aged ≥65, and predominantly White (48.6%) or Asian (31%) ethnic groups. The area is a mixed urban, semi-urban and rural area of 267.8KM2.

#### 5.3.2 Unique service users for new patient assessment

The number of unique service users for new patient assessment was 2,557 in 2022/23, 2,892 in 2023/24 and 2,868 in 2024/25.

#### 5.3.3 Time spent on all nursing and palliative and end-of-life care activities

The total number of hours allocated for all nursing activities was 389,625 in 2022/23, 368,917 in 2023/24 and 355,024 in 2024/25. The total hours allocated to all nursing activities was on average 370,188 per year.

The total hours allocated to palliative and end-of-life care activities was 35,632 in 2022/23, 36,078 in 2023/24 and 34,362 in 2024/25. The total hours allocated to palliative and end-of-life care activities was on average 35,357 per year.

Overall, palliative and end-of-life care represents approximately 9.6% of all community health nursing activities. By activity, most time was spent on reviewing and supporting patients (57%), followed by syringe driver administration (14%), new patient assessments (13%), symptom control reviews (10%), crisis management (4%), bereavement support visits (2%) and care of the deceased (1%). See Table 3 in Supplementary File 1 for summary of hours spent by activity by year.

#### 5.3.4 Comparing time spent on all nursing care and palliative and end-of-life care per 1,000 population by year

The total number of hours spent on all nursing care is reducing year-on-year, from 338 hours per 1,000 population in 2022/23, to 322 hours in 2023/24, and 310 hours in 2024/25. In contrast, the total time spent on palliative and end-of-life care activities remains stable year-on-year at 31 hours per 1,000 population in 2022/23, 31.5 in 2023/24, and 30 in 2024/25.

#### 5.3.5 Percentage of all nursing care time that was spent on palliative and end-of-life care in and out-of-hours by year

In the context of all care provided, Figure 7 suggests that more time is spent on out-of-hours palliative and end-of-life care in comparison to in-hours. It also shows that the time spent on in-hours palliative and end-of-life care remained stable across the three years. In contrast, out-of-hours activity was higher in 2022/23 and 2023/24 before decreasing to 14% in 2024/25.

**Figure 6.**
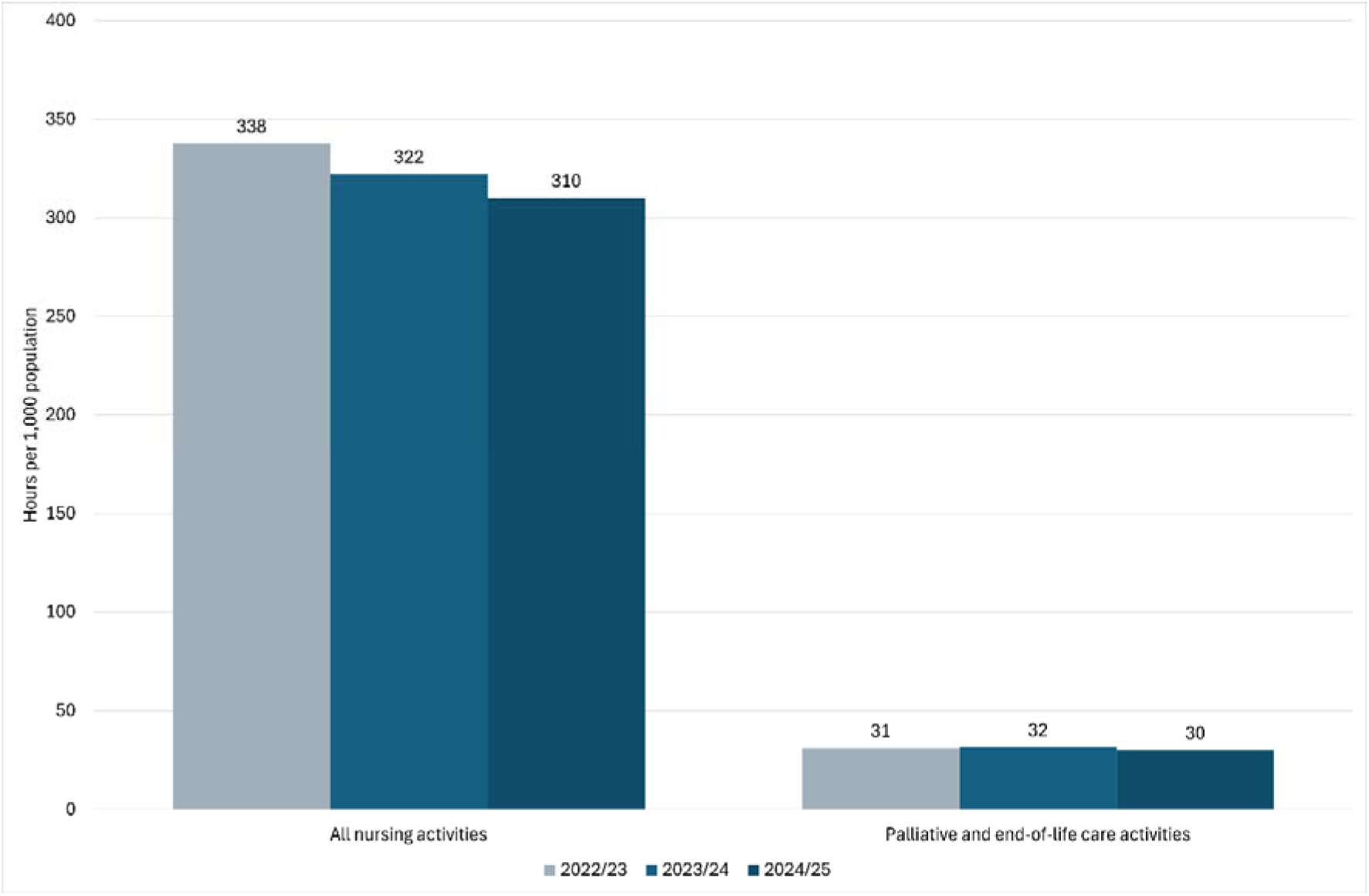
The hours spent on all nursing care and palliative and end-of-life care per 1,000 population by year

**Figure 7.**
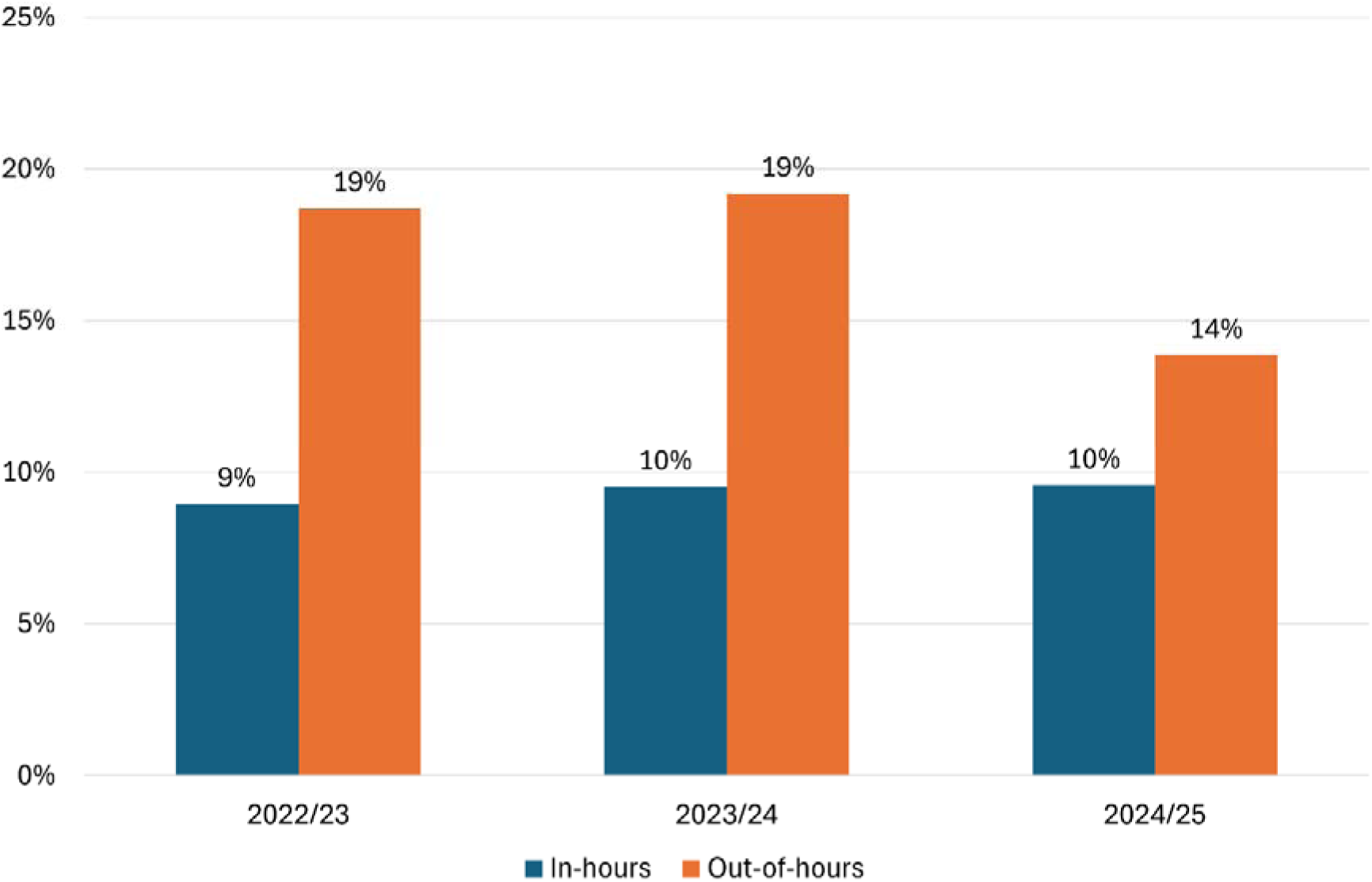
The percentage of all nursing care time that was spent on palliative and end-of-life care in and out-of-hours by year

#### 5.3.6 Comparing the percentage of time spent in and out-of-hours on palliative and end-of-life care by activity and year

In comparing in and out-of-hours, new patient assessments, personal care at the end of life, syringe driver administration, symptom control review, and bereavement support visits appeared to be more commonplace in-hours. Whilst crisis management and care of the deceased commonly occurred out-of-hours.

#### 5.3.7 Minutes per unique service user by palliative and end-of-life care activity over years

Figure 9 shows the number of minutes spent per unique service user for each palliative and end-of-life care activity by in and out-of-hours and year (see Table 3 and Table 4 in Supplementary File 1 for hours spent and number of unique service users by activity and year). The time spent on new patient assessments per unique service user has remained stable at approximately 90 minutes per service user. For review and support, the time spent in minutes has increased out-of-hours per service user (from around 1 hour to 1 hour 15 minutes per service user), whilst in hours has remained steady at about 30 minutes per service user. The time spent in minute on symptom control review have increased in the past couple of year in-hours (from under 1 hour to almost 1 hour and 15 minutes per service user) but has remained consistent out-of-hours (i.e. about 45 minutes per service user).

**Figure 8.**
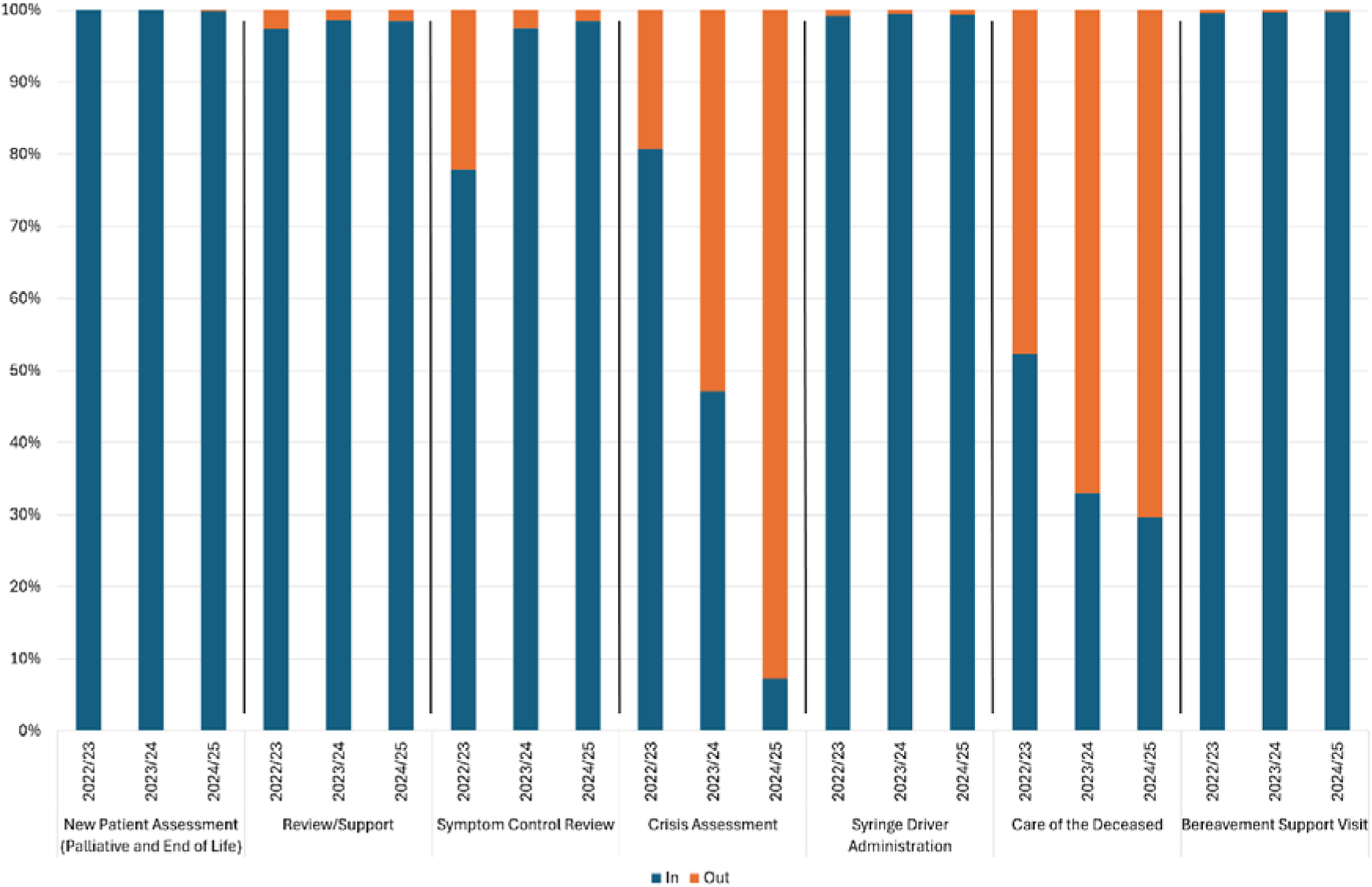
Comparing the percentage of time spent on palliative and end-of-life care in and out-of-hours by activity and year

**Figure 9.**
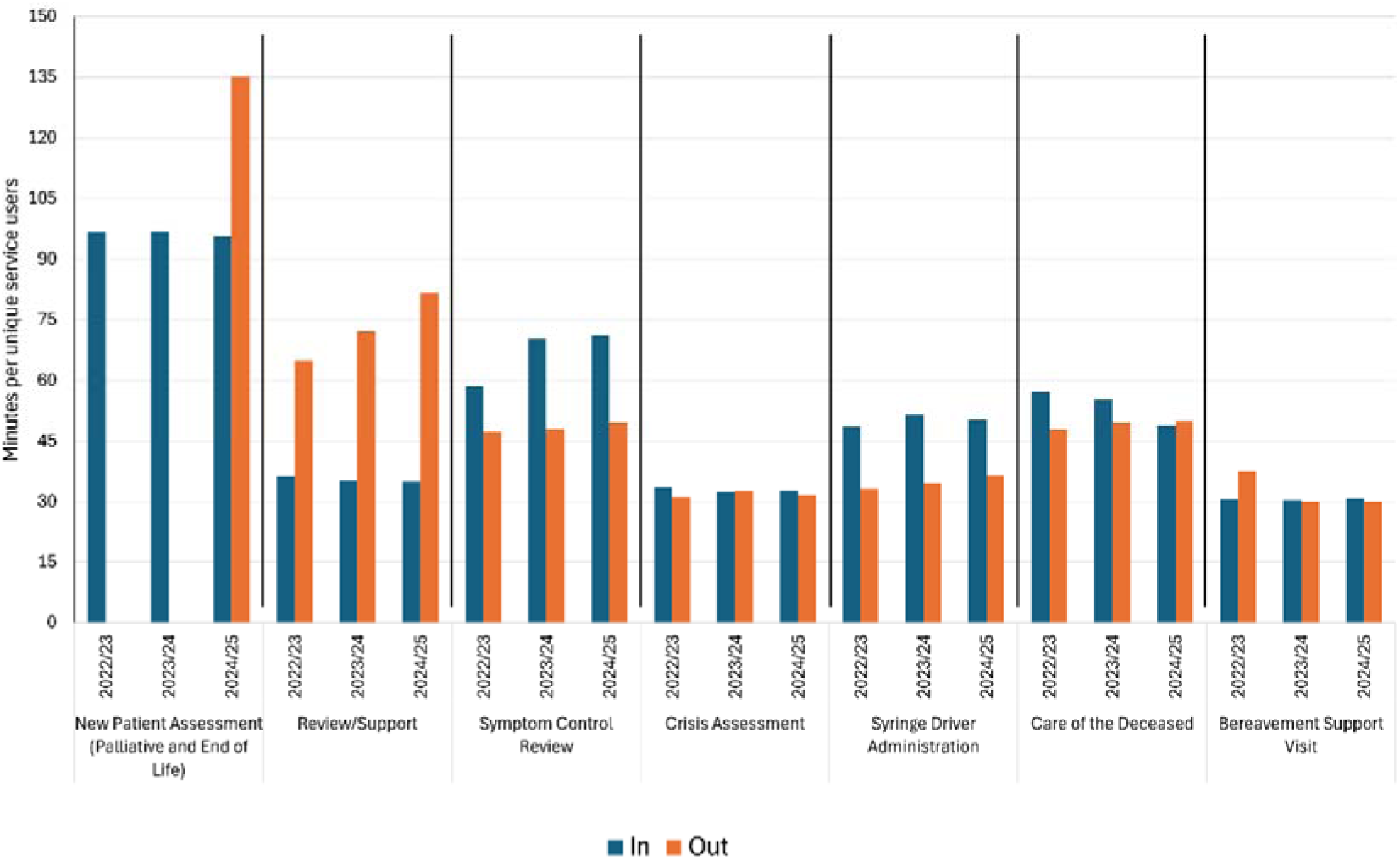
Minutes per unique service user for each palliative and end-of-life care activity by in and out-of-hours Note: the ‘N’ varies per activity and in and out-of-hours.

The time spent in minutes has remained stable across years and is similar in and out-of-hours for crisis assessment and bereavement support visits (taking approximately 30 minutes per unique service user). More time is spent per service user on syringe driver administration per service user in hours compared to out-of-hours

## 6. Discussion

This work provides a novel view of the scale and nature of palliative and end-of-life care provision by community health nursing services, drawing together a wider view from nationally collected data and a more detailed task-based regional dataset. It shows that there is a marked increase in referrals to community health nursing services, but a reduction in time on caseload, alongside a stable number of unique service users being seen in year.

There has been a slight increase in the average number of contacts per 1,000 weighted population, particularly since 2022. The length of each contact (in minutes) has remained steady, but the days spent on caseload for each referral is falling significantly. The time spent of palliative and end-of-life care activities has been consistent despite the time spent on all nursing care falling year on year. Palliative and end-of-life care (measured conservatively) accounts for 9.6% of all community nursing clinical time and commonly occurs out-of-hours (particularly with crisis management and care of deceased).

Together, these findings indicate that models of community health nursing care are changing; from longer-term and sustained relational care toward shorter episodes of care. Projected estimates indicate that the number of annual deaths and the proportion occurring in the community will continue to increase over the coming decades.^6,7^ In line with this, we expected to see an increase in the number of unique service users year by year. However, the overall number of unique service users has remained stable. The increasing number of referrals (as opposed to service users, which remains stable), and declining time on caseload, signals notably shorter episodes of care followed by re-referrals when need increases again. It is unclear from the data whether patients are being re-referred for the same problem or new ones. Although shorter episodes of care do not necessarily indicate lesser experiences of care, it may indicate that patients are experiencing poorer continuity of care. In addition, we do not know if this masks unmet population need, or possibly that referrals for palliative and end-of-life care are now occurring closer to death. We do know that community health nurses’ workload is increasing due to changes in ways of working by the wider multi-disciplinary team and extending their roles in palliative and end-of-life care.^8,11^ In addition, challenges with accessing other community-based professionals (such as general practitioners) means that community health nursing teams are heavily relied upon.^28^ These challenges are noted to result in moral distress and an impaired capacity to provide good clinical care, which may further impact the retention of nurses within an already shrinking workforce.^8,9^ It is also time-consuming and stressful for people with advanced illness and their families to navigate and manage different services.^28^

The proportion of time allocated to palliative and end-of-life care in the regional dataset adds a contemporary estimate to earlier heterogenous values. It is (conservatively) estimated that palliative and end-of-life care accounts for 9.6% of all community health nursing clinical time, and highlights that a large proportion of this occurs outside of normal working hours. Other estimates have ranged from 6% of clinical time on palliative care^13^ to spending 23.5% of their most recent shift providing end-of-life care.^15^ Differences across estimates are likely to arise from definitions and measurement approaches and highlights the need for a more standardised approach. There are also issues with the community nursing workforce identifying palliative and end-of-life care needs,^4^ which will impact on how these activities are recorded in acuity tools. Palliative and end-of-life care needs may be recognised late on, such as the last weeks or days of life. The high proportion of palliative and end-of-life care occurring out-of-hours demonstrates the imperative for 24/7 provision.^29,30^

One aspect we could not identify within either dataset was how many deferred appointments related to palliative and end-of-life care. Literature has indicated community health nurses prioritise palliative and end-of-life care.^1^ It could potentially be inferred from the regional data that this is the case, as time spent on palliative and end-of-life care has remained stable across years even when total care hours have decreased. Although, a cross-sectional survey has demonstrated that the distribution of missed care was spread evenly across all types of nursing care (including aspects of palliative and end-of-life care).^31^ Further understanding is needed around this.

Less contact with professionals than desired in the community was noted in a nationally representative survey of bereaved family carers in England and Wales.^28^ Based on the Queen’s Institute of Community Nursing (QICN),^32^ community health nursing visits should be a minimum of 30 minutes (excluding travel time). In the national dataset, the average length of contacts (for all general nursing care) across years were mostly just under 30 minutes. There may be a limit to any reduction to contact time, where it is not possible to be any quicker. In contrast, the regional dataset indicated the recommended time was met per unique service user or were greater (depending on palliative and end-of-life care task). The increase in crisis assessment occurring out-of-hours in our data potentially signals the constraints of care provided in-hours. Other research has also highlighted out-of-hours care as a barometer for proactive in-hours care.^4,29^

### 6.1 Patient and public involvement perspectives

Our Patient and Public Involvement Group reflected on the data and noted the following:

- Data contributions:

`o` There is limited transparency around how data metrics are chosen and why. Patients, family carers and members of the public should be involved in deciding what data is gathered and how it is then interpreted.
- Limitations with the data:

`o` It is important to understand what happens between referral and contact for people with palliative and end-of-life care needs, including how they are identified and how long the wait times are.
`o` Further granularity is needed with some data groupings, such as what ‘review and support’ accounts for in the regional dataset.
`o` Task-based approaches to data overlook holistic care and patient experience.

### 6.2 Strengths and limitations of the work

To our knowledge, this is the first time that national and regional datasets have been used to describe community health nursing in palliative and end-of-life care. Data on specialist palliative and end-of-life care are reported,^33^ but we have limited data on community health nursing. However, it is critical to understand given the projected demographic challenges and policy focus on shifting care from hospitals to the community.^6,7,16,17^ Combining these datasets increases our insights into the scale and nature of palliative and end-of-life care provided by community health nursing services.

One limitation is that this was a secondary analysis of existing data, which meant that there was no control over what data is collected, the variables and the level of missing data. For example, we were unable to determine whether the increase in referrals was due patients to being re-referred for the same or different problems and had limited information around deferred care. The data on palliative and end-of-life care at a national level was more limited, and we recognise that how palliative and end-of-life care is defined may also vary by service. More detail on palliative and end-of-life care activities and the time spent on these were available in the regional dataset. Although, there was likely underreporting of urgent/unplanned care, which means our estimates will be conservative.

### 6.3 Implications for policy and practice

Models of community health nursing care appear to be changing, and it is important to understand the implications of this. Standardised longitudinal data collection on community health nursing in palliative and end-of-life care is essential to achieve policy ambitions of moving more care to the community setting. Capturing core palliative and end-of-life care tasks (and co-designing this with service users) will support understanding what workforce is needed, enable robust benchmarking and help to understand whether provision is equitable.

Changing demographics and population demand mean that planning ahead to ensure sufficient provision and workforce in community health nursing services is crucial. We need better data, including consistent data over time and more evidence on time spent on different palliative and end-of-life care activities, how much of this care is being deferred (including for how long and why), and how care is meeting the needs of those using the service. In addition, the high proportion of care occurring outside of normal working hours in the regional data highlights the imperative for 24/7 provision.

## 7. Conclusions

Increasing referrals to community health nursing services, the stable number of unique service users seen in the year, and shorter time on caseload indicate a system under pressure and a shift towards shorter episodes of care. Despite this picture from national data, the time spent on palliative and end-of-life care activities by community health nursing teams remains stable and conservatively accounts for 9.6% of all nursing care provided. A high proportion of this care occurs out-of-hours, with a noticeable increase in crisis management. Aligning the workforce, skill mix and out-of-hours provision with the challenges raised, while improving standardised recording of palliative and end-of-life care, is essential to support high-quality person-centred care at home. In addition, understanding the scale and nature of community health nursing in palliative and end-of-life care combined with an understanding of what matters to people receiving the care, will help to guide workforce planning, service configuration and quality of care.

## Data Availability

All data produced in the present study are available upon reasonable request to third party contacts.

## Notes

### Competing Interest Statement

The authors have declared no competing interest.

### Funding Statement

This research is funded through the NIHR Policy Research Unit in Palliative and End of Life Care, reference NIHR206122. FEMM is a UK National Institute for Health and Care Research (NIHR) Senior Investigator. The views expressed are those of the author(s) and not necessarily those of the NIHR or the Department of Health and Social Care.

### Summary of Updates

Additional details needed to be added within the 'Ethical considerations' section. 'The National Health Service (NHS) Benchmarking Team' should read as 'The National Health Service (NHS) Benchmarking Network Delivery Team'. In addition, 'hosted by the England and Community Health and Care NHS Trust' should read as 'hosted by East of England Community Health and Care NHS Trust'.

