## Supplementary file 1 for "Contributions to palliative and end-of-life care by community health nursing services: improving care through national and regional service evaluations"

Supplementary file 1 Supporting information

### Methods

#### NHS Benchmarking Data sources

Table 1 Breakdown of Trusts and submissions in England by year

| **Year** | **Trusts** | **Submissions** |
| --- | --- | --- |
| 2013 | 63 | 69 |
| 2014 | 45 | 52 |
| 2015 | 47 | 52 |
| 2016 | 53 | 69 |
| 2017 | 50 | 62 |
| 2018 | 47 | 65 |
| 2019 | 55 | 75 |
| 2020 | 40 | 45 |
| 2021 | 46 | 59 |
| 2022 | 50 | 63 |
| 2023 | 51 | 75 |
| 2024 | 48 | 65 |

#### NHS Benchmarking Metrics and Definitions

- Population served (the population figure which this service is commissioned):
  - Actual: based on Office for National Statistics resident population.
  - Weighted: Base populations were weighted by demography (age and sex) and GP practices needs index (including deprivation) to reflect local health and care needs.^1^
- Workforce was considered in terms of whole time equivalent in establishment and whole time equivalent in post:
  - In establishment: the total number of budgeted positions, including both occupied roles and vacancies.
  - In post: the actual number of employees occupying roles.
- Referrals received and accepted, and sources of referrals.
- Number of unique service users seen in the year.
- Contacts (face-to-face, non-face-to-face (i.e. telephone/video call) and total) and average length of contacts.
- Average time on caseload (in days).
- Location of care and types of care provided, and support to other teams/processes.
  - Location of care: Services reported various locations in which they provided their services. Some of these locations were grouped due to variations over time. Groupings are indicated beneath the relevant tables.
  - Types of care provided: Several types of care were listed; care that was explicitly related to palliative and end-of-life care were included alongside other aspects of care that were deemed related but may also form part of general nursing care (including ‘pain control’, ‘medication prescribing, administration and review’, ‘continuing healthcare assessment’, ‘percutaneous endoscopic gastrostomy (PEG) feeding’, and ‘nasogastric tube feeding’). This variable indicated only whether services provided this type of care or not.
- Deferred care defined as a visit that was planned to take place that day, but has been postponed to another day, as staff capacity did not meet demand and prioritisation of patients was required.

#### Care in Focus – Community health nursing team structure

Each locality has a Service Manager (Operational Band 8a) and a Quality Matron (Band 8a) and comprises four planned teams and one unplanned team. Figure 1 illustrates the team structure. For out-of-hours, two teams (evening service and night service) have a similar team structure as outlined below but cover all localities.

Clinical Operational Team Manager - Band 7 x 1 WTE

Deputy Team Leader - Band 6 x 4 WTE

Triage Nurse - Band 6 x 1 WTE

Community Staff Nurse - Band 5 x 6 WTE

Nurse Associate - Band 4 x 6 WTE

Healthcare Assistant - Band 3 x 3 WTE

Figure 1 Team structure for Birmingham community nursing services

#### Care in Focus - List of palliative and end-of-life care activities and groupings

From a comprehensive list of activities, those relating to palliative and end-of-life care were selected. There were some categories that have been introduced and allocation to these varied, we therefore grouped these categories to account for these challenges. In addition, community nurses can struggle to recognise palliative and end-of-life care needs; these were grouped appropriately to account for this.

Table 2 Groupings for palliative and end-of-life care activities within the 'Care in Focus' regional dataset

| **Original Label** | **Grouping** |
| --- | --- |
| Assessment – New Patient (End-of-Life Care) | Assessment |
| Assessment – New Patient (Palliative) |  |
| End-of-Life Care – Support/Review (Last Days/Weeks of Life) | Review/Support |
| Palliative – End-of-Life Care (Personal Care) |  |
| Palliative – Support |  |
| Palliative – Support/Review (Last Months of Life) |  |
| Palliative – Support/Review (Last 12 Months of Life) |  |
| Palliative – Symptom Control Review | Symptom Control Review |
| Urgent Community Response Palliative – Crisis Assessment | Crisis Assessment |
| Medication – Administration Syringe Driver | Syringe Driver Administration |
| End-of-Life Care – Care of the Deceased | Care of the Deceased |
| Palliative – Verification of Death |  |
| Palliative – Bereavement Support Visit | Bereavement Support Visit |

### Results

#### National Cross-sectional Analysis (2024): Sources of referrals

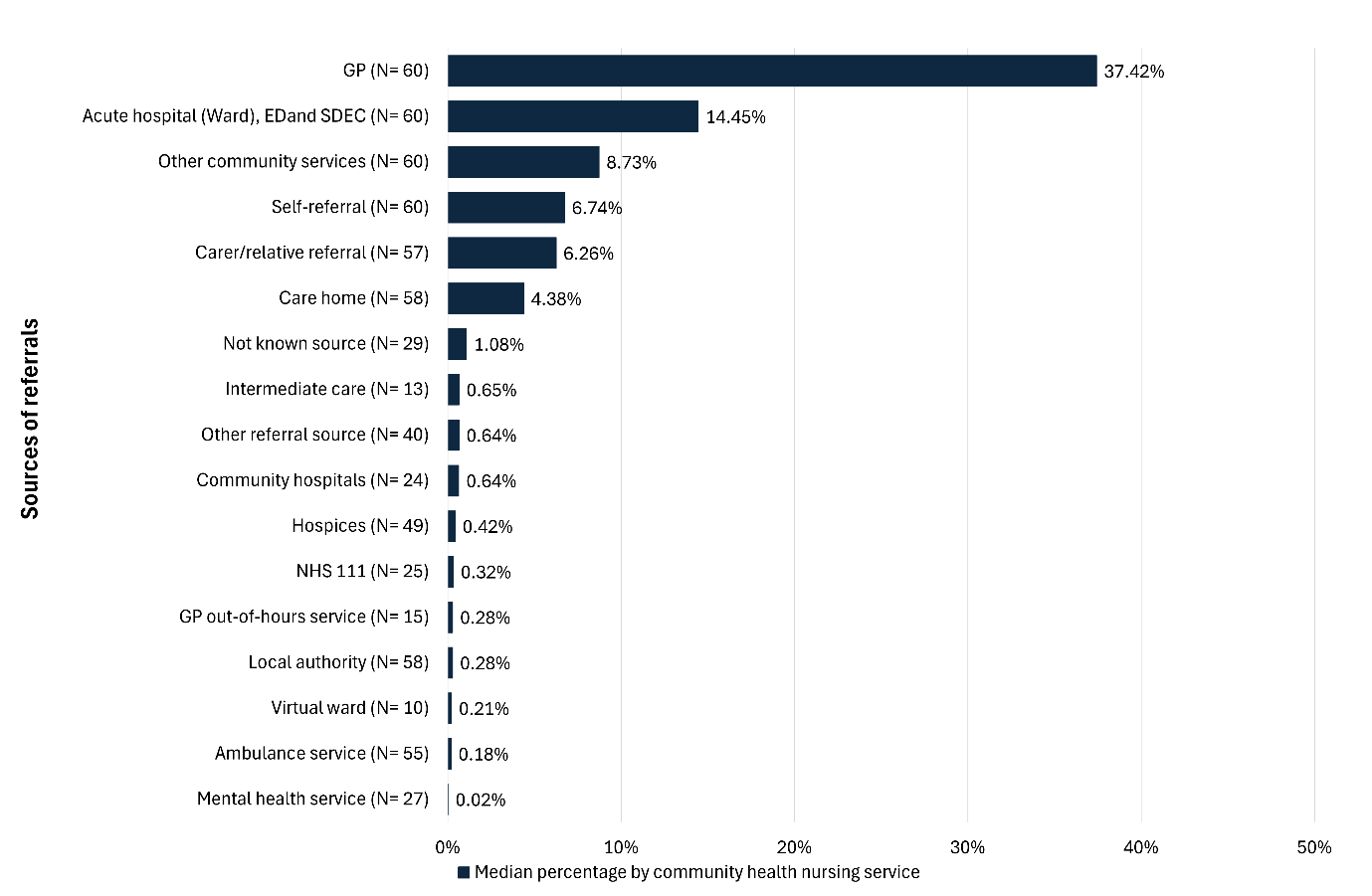

Note: ED: Emergency Department, GP: General Practice, SDEC: Same Day Emergency Care

Figure 2 The median percentage for sources of referral to community health nursing, by community health nursing service

#### National Cross-sectional Analysis (2024): Support to other teams and processes

Community health nursing services provide support to other community services (including teams and processes), which is presented in Table 3.

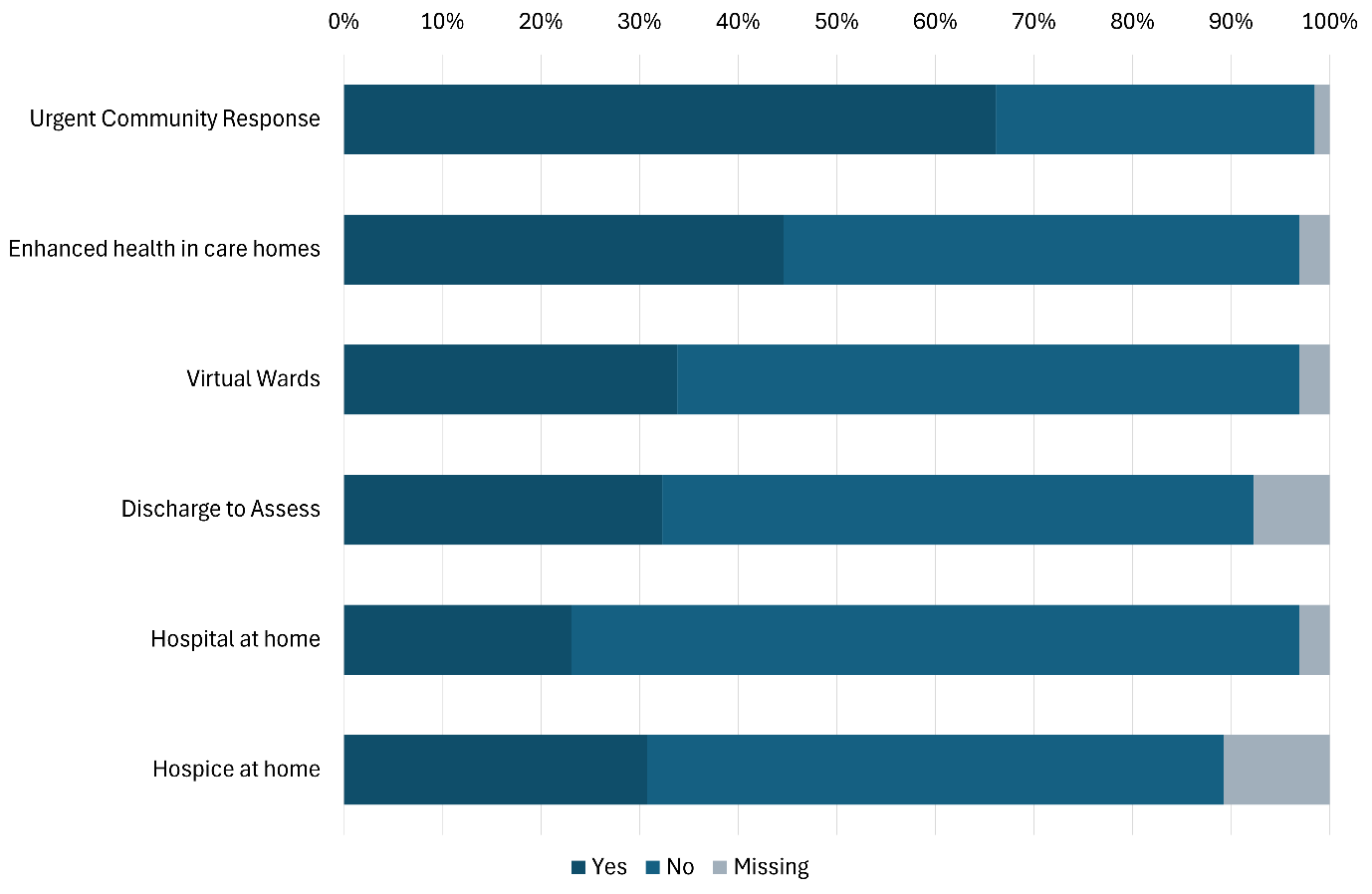

Figure 3 Support provided by community nursing teams to other teams or processes (n= 65)

#### Regional Repeated Cross-sectional Analyses: Unique service user by palliative and end-of-life care by activity and year

Table 3 Summary of the hours spent on palliative and end-of-life care activities by year

| **Activity** | **2022/23** | **2023/24** | **2024/25** | **Total** |
| --- | --- | --- | --- | --- |
| New patient assessment | 4,118 | 4,665 | 4,574 | 13,356 |
| Review/support | 21,152 | 19,799 | 19,381 | 60,332 |
| Symptom control | 4,436 | 3,161 | 3,044 | 10,640 |
| Crisis management | 1,315 | 2,552 | 812 | 4,678 |
| Syringe driver administration | 3,683 | 5,074 | 5,932 | 14,689 |
| Care of the deceased | 308 | 257 | 129 | 695 |
| Bereavement support visit | 621 | 571 | 492 | 1,684 |
| Total | 35,632 | 36,078 | 34,362 | 106,072 |

Table 4 Number of unique service users by palliative and end-of-life care activity and year

| **Palliative and end-of-life care activity** | **2022/23** | **2023/24** | **2024/25** |
| --- | --- | --- | --- |
| New Patient Assessment (Palliative and End of Life) | 2,557 | 2,892 | 2,868 |
| Review/Support | 34,587 | 33,667 | 32,951 |
| Symptom Control Review | 4,791 | 2,729 | 2,585 |
| Crisis Assessment | 2,393 | 4,717 | 1,533 |
| Syringe Driver Administration | 4,574 | 5,936 | 7,115 |
| Care of the Deceased | 353 | 301 | 156 |
| Palliative - Bereavement Support Visit | 1,216 | 1,127 | 960 |

### References

1.	NHS England. Community Services allocations formula: For 2019/20 to 2023/24 revenue allocations. NHS England; 2019.
